# Effects of changes in calculating GFR using KDIGO standards: Discordance in the Staging and Timing of Diagnosis of Chronic Kidney Disease

**DOI:** 10.1101/2023.12.21.23300415

**Authors:** Charlotte Baker, Samuel Gratzl, Patricia J Rodriguez, Michael Simonov, Brianna M Goodwin Cartwright, Rajdeep Brar, Nicholas L Stucky

## Abstract

**Introduction:** Chronic kidney disease (CKD) is a highly prevalent disease with disparities in diagnosis and treatment. Until recently, primary diagnosis for CKD was based on equations that incorporated race and have demonstrated racial bias. This study had two aims comparing outcomes for Black patients and their counterparts: 1) whether using the new 2021 CKD-EPI equation led to decreased disparity with time to diagnosis; and 2) whether there was discordance in the staging between the two equations at potential diagnosis point.

**Methods:** We evaluated patients aged 18 and over with non-hospitalization related serum creatinine laboratory results between January 1, 2016 and September 30, 2023. We estimated the GFR for each patient using the 2009 and 2021 CKD-EPI creatinine equations. We assessed stage discordance for stages 3a, 3b, 4, and 5 using chi-square tests and the Cochran-Mantel-Haenszel. We used multivariate logistic regression to assess a change in staging based on the equation used.

**Results:** 15.5% of the 8,080,889 patients included in this study were Black. The median age was 57 years and 15.3% of patients met the criteria for stage 3a CKD or higher using either equation. Discordance in staging by equation and by race existed, especially for Black patients at stages 3a and 3b. 40% of Black patients identified as stage 4 using the 2021 equation were 3b or lower using the 2009 equation.

**Discussion:** Well established medical algorithms with race components are being re-examined. We found more disparity with the initial staging of the disease. The disconnect in the timing of staging by equation for Black patients means a number of these patients may not have received the appropriate treatment. Future work should elucidate the impact of the change in CKD staging with the 2021 CKD-EPI creatinine equation on treatment.

**Conclusion:** Significant disparity exists in the timing and staging of CKD by CKD-EPI equation and by race.

## Introduction

Chronic kidney disease (CKD) is a highly prevalent disease^1^ and is associated with common co-morbidities such as hypertension and diabetes.^2^ Additionally there are disparities in diagnosis and treatment of this disease.^3^ Of the 14% of adults in the US with CKD, as many as 9 in 10 people do not know they have it.^4^ Black and Hispanic patients are more likely to develop end stage disease than white patients and are also less likely to be able to obtain care that slows the progression of CKD.^5^ More than 35% of patients on hemodialysis are Black and patients that are Black are less likely than their counterparts to receive a transplant evaluation.^6, 7^ Compounding these disparities, until recently primary diagnosis for CKD was based on equations that incorporated race and have demonstrated racial bias.^8, 9^ The inclusion of a race adjustment in the equations for CKD were based on the assumption that Black people have a higher muscle mass, than other racial groups^9^ and therefore should have a higher baseline serum creatinine level.^10^ In practice, the Chronic Kidney Disease Epidemiology Collaboration (CKD-EPI) 2009 GFR estimation equation included a multiplier of −1.157 that resulted in a lower GFR calculation (indicating higher kidney function) in Black patients than in other racial groups with similar input values for creatinine. The National Kidney Foundation and American Society of Nephrology (NKF-ASN) Task Force recommended changes to the determination for GFR in the United States; “[t]here is no evidentiary basis for keeping a race coefficient in the eGFR calculation”) or to reinforce racial bias by including race in the calculation but remove it during clinical reporting of GFR.^9^ In the last few years, after a push by knowledgeable medical students^11, 12^ and experts, the race adjustment has been removed and GFR equations have been updated. However, the effect of using these equations in clinical practice is still being understood.^13^ Some studies have proposed that the new equation leads to inaccurate GFR calculations in transplant patients, racial bias in outcomes such as kidney failure and mortality, and still needs further evaluation to assess accuracy in Black adults.^14, 15, 16^ Other studies have found that the new equation is advantageous to patients of color and the elderly as they have the potential to be identified earlier and receive more equitable access to care.^17^ We aimed to understand whether using the new 2021 CKD-EPI equation led to decreased disparity between Black patients and their counterparts with regards to how quickly they should have been diagnosed with CKD. We also aimed to find whether there was discordance in the staging between the two equations at potential diagnosis for Black patients compared to their counterparts.

## Methods

### Study Data

This study used a subset of Truveta Data. Truveta (https://www.truveta.com/) provides access to continuously updated and linked electronic health record (EHR) from a collective of US health care systems, including structured information on demographics, encounters, diagnoses, vital signs (e.g., weight, BMI, blood pressure), medication requests (prescriptions), medication administration, laboratory and diagnostic tests and results (e.g., serum creatinine tests and values), and procedures. Updated EHR data are provided daily to Truveta by constituent health care systems. In addition to EHR data for care delivered within Truveta constituent health care systems, and social determinants of health (SDOH) information are made available through linked third-party data. SDOH data include individual-level factors, including income and education. Data are normalized into a common data model through syntactic and semantic normalization. Truveta Data are then de-identified by expert determination under the HIPAA Privacy Rule. Once de-identified, data are available for analysis in R or Python using Truveta Studio. Data for this study were accessed on December 14, 2023.

### Eligibility criteria

We evaluated patients with non-hospitalization related serum creatinine laboratory test values between January 1, 2016 and September 30, 2023 using a subset of Truveta Data. Our population included patients that were 18 years or older at the time of the first laboratory test within the study period. We were interested in identifying all patients that would meet the criteria for CKD based on their GFR estimation calculation so patients did not need a billing or diagnosis code for CKD to be included, just a history of serum creatinine laboratory results. Patients were required to be male or female sex and to have non-missing or unknown race. We combined all non-Black or African-American race categories into a “Not Black or African-American” category for the analysis to be in alignment with the categories used in the 2009 CKD-EPI GFR equation. Patients must have had no fewer than three serum creatinine lab values spanning a period of greater than 90 days. We sought to identify the point in time when patients could have been diagnosed with CKD (GFR < 60 for a period of 60 or more days) using both the 2009 CKD-EPI creatinine equation and the 2021 CKD-EPI creatinine equation.

We estimated the GFR for each patient using both equations (Table 1). Serum creatinine laboratory tests and results were identified using LOINC codes 14682-9, 2160-0, 35203-9, 38483-4,

**Table 1:**
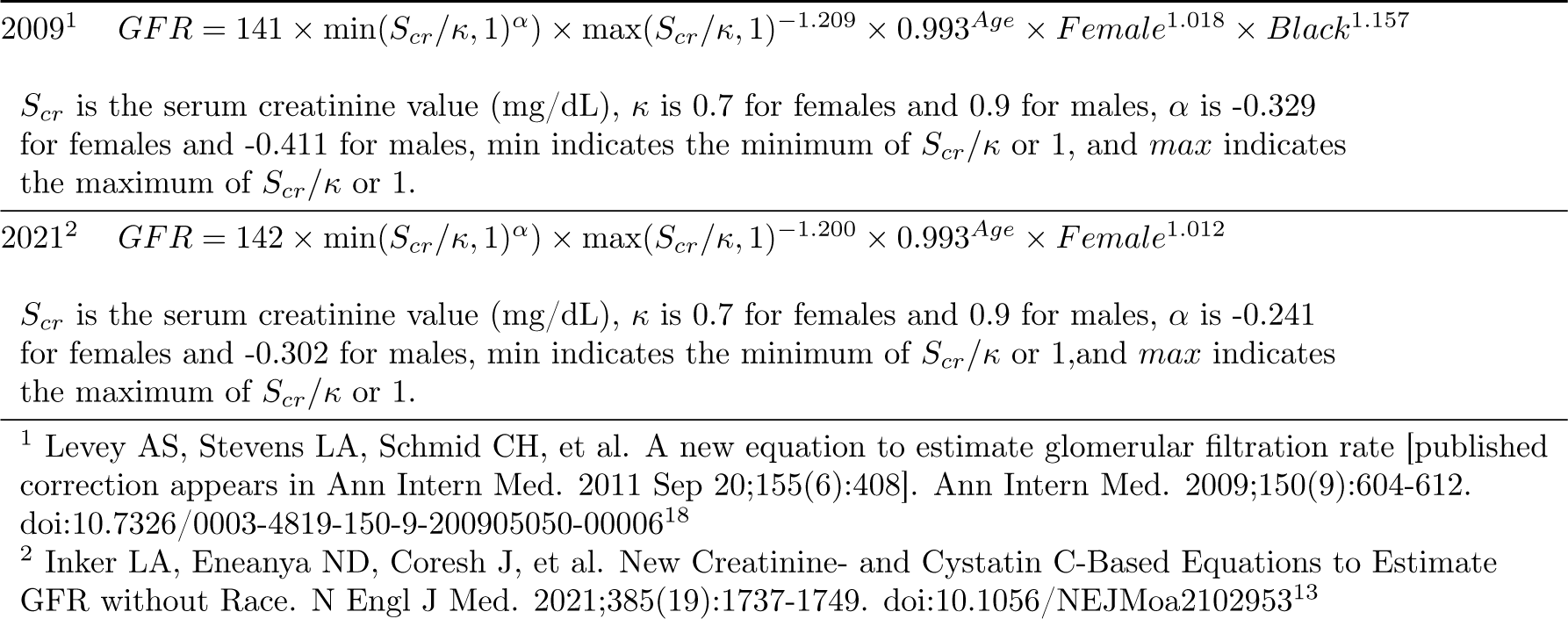
2009 and 2021 Chronic Kidney Disease Epidemiology Collaboration (CKD-EPI) Creatinine Equations for Estimating Glomerular Filtration Rate.

44784-7, 59826-8, and 77140-2.

Patients could not be pregnant or have a SNOMED CT, ICD-9-CM, and ICD-10-CM billing or diagnosis code for polycystic kidney disease or acute kidney injury during the study period (Supplement).

### Outcome

This study focused on two specific outcomes - 1) discordance in staging of CKD (stages 3a, 3b, 4, or 5) between the two equations; and 2) the odds of identifying CKD using the 2021 equation earlier compared to the 2009 equation.

### Covariates

We included demographics, comorbidity, and SDOH factors in this analysis. Because of their importance in the equations, we included age, sex, and race. We included ethnicity because of known disparity of CKD for Hispanic patients. We categorized age into three categories (18-44, 45-64, and 65 and over) and as a continuous variable for describing the data. We used the continuous value of age in statistical modeling.

To adjust for comorbidity, we included hyperkalemia (a frequent co-occurring condition in patients with CKD) and used the Elixhauser Comorbidity Index^19^ with van Walraven weights.^20^ Similar to existing literature,^21^ we created a three level variable for the interpretation of the Elixhauser Comorbidity Index. Scores of less than 11 were categorized as “low comorbidity”, 11 to 15 were categorized as “medium comorbidity”, and greater than 15 were categorized as “high comorbidity”. We included four SDOH variables: combined household annual income range (9 categorical increments from $0 - 25,000 to $100,000 and above); number of people in the household, an indicator of how much resources such as time and finances may impact time to dedicate to health needs; and distance to family or other close ties such as friends or acquaintances as an indicator of proximity to a support network. We also included marital status, a known epidemiological indicator of obtaining medical care and improved health outcomes.

### Statistics

We estimated the GFR using the 2009 and 2021 CKD-EPI creatinine equations for each available serum creatinine value (Table 1). As patients had multiple serum creatinine laboratory results across multiple days, we repeated an iterative process with each equation to identify the first pair of visits (initial visit and subsequent visit) at least 60 days and no more than 365 days apart where both GFR values were less than 60 in order to classify someone as having CKD. We used the GFR value to classify the patient’s stage of CKD^22^ at the identified first visit in the pair and a stage of CKD at the second identified visit in the pair. This resulted in two pairs of visits (one for the 2009 equation and one for the 2021 equation) and subsequently two GFR values (one using the 2009 equation and one using the 2021 equation). If a patient was not classified as having Stage 3a (GFR between and inclusive of 45 and 59) or worse (GFR less than 45) CKD using either equation, they were marked as not having CKD for the purposes of this study.

### Sub-analysis for patients with CKD

Among the subgroup of patients with a stage of 3a-5 CKD diagnosis using either formula, we conducted three separate analyses.

First, we assessed discordance of staging between the 2021 equation and the 2009 equation. Differences were evaluated using chi-square tests. We further stratified these results by race and used the cochran mantel haenszel test to determine if the amount of discordance was greater for Black or African-American patients.

Using the two pairs of visits (one for the 2009 equation and one for the 2021 equation) from the iterative process outlined above, we identified which equation identified the patient’s CKD status earlier. We made a three level variable with the result showing if first with 2009 equation, if first with 2021 equation, and then if no difference.

Finally, we used multivariate logistic regression to assess whether there was an increased chance to have a higher (denoting more severe disease) or lower (denoting less severe disease) initial CKD stage using the 2021 equation, compared to the 2009 equation. All covariates were included in the original full model. Patients with missing values for SDOH or other covariates were excluded from the analysis. We used backwards model selection to create the best fit model and compared models using the AIC.

### Stats program and packages used

We used Python to clean the data and R for statistics. Data preparation was completed using Python 3.10.12^23, 24^ and packages pandas,^25, 26^ NumPy,^27^ PyYAML,^28^ and matplotlib.^29^ All analyses were conducted in R Version 4.2.3^30^ using packages dplyr,^31, 32^ ggplot2,^33, 34^ gtsummary,^35^ broom,^36^ MASS,^37^ xtable,^38^ reshape2,^39, 40^ readr,^41^ arrow,^42^ stringr,^43^ sparklyr,^44^ and SparkR.^45^

## Results

### Description of study patients

8,080,889 patients with serum creatinine laboratory results were included in the study (Table 2). 15.5% were Black or African American [henceforth referred to as Black]. The median age for Black patients was 51 years (IQR 36, 64) and the median age for non-Black patients was 58 years (IQR 43, 70). The population was 58% female. 1,239,185 (15.3%) patients met the criteria for CKD stages 3a, 3b, 4, or 5 using either the 2009 (94.9%) or 2021 (98.6%) CKD-EPI serum creatinine only equation.

**Table 2:** Demographic and comorbidity characteristics of people with serum creatinine laboratory tests after January 1, 2016 in a subset of Truveta data.

| Characteristic | Black*,<br>N = 1,253,042<sup>1</sup> | Not Black*,<br>N = 6,828,038<sup>1</sup> | Overall,<br>N = 8,081,080<sup>1</sup> |
| --- | --- | --- | --- |
| Age Group |  |  |  |
| 18-44 | 483,411 (39%) | 1,891,386 (28%) | 2,374,797 (29%) |
| 45-64 | 482,972 (39%) | 2,482,725 (36%) | 2,965,697 (37%) |
| 65+ | 286,626 (23%) | 2,453,769 (36%) | 2,740,395 (34%) |
| Age (Median (IQR)) | 51 (36, 64) | 58 (43, 70) | 57 (42, 69) |
| Sex |  |  |  |
| Female | 806,667 (64%) | 3,918,090 (57%) | 4,724,757 (58%) |
| Male | 446,342 (36%) | 2,909,790 (43%) | 3,356,132 (42%) |
| Ethnicity |  |  |  |
| Hispanic or Latino | 12,437 (1.1%) | 734,091 (11%) | 746,528 (9.8%) |
| Not Hispanic or Latino | 1,168,387 (99%) | 5,724,669 (89%) | 6,893,056 (90%) |
| Marital Status |  |  |  |
| Unmarried and Never Married | 527,742 (49%) | 1,462,852 (24%) | 1,990,594 (28%) |
| Domestic Partner and Civil Union | 50,142 (4.6%) | 457,713 (7.5%) | 507,855 (7.1%) |
| Married | 317,585 (29%) | 2,991,688 (49%) | 3,309,273 (46%) |
| Divorced | 85,478 (7.9%) | 506,089 (8.3%) | 591,567 (8.2%) |
| Other | 16,127 (1.5%) | 47,284 (0.8%) | 63,411 (0.9%) |
| Widowed | 81,548 (7.6%) | 632,680 (10%) | 714,228 (10.0%) |
| Number of People in the Household |  |  |  |
| 1 | 461,615 (41%) | 1,810,709 (30%) | 2,272,324 (32%) |
| 2 | 221,705 (20%) | 1,496,066 (25%) | 1,717,771 (24%) |
| 3 | 152,952 (14%) | 1,053,771 (17%) | 1,206,723 (17%) |
| 4 | 105,919 (9.4%) | 703,911 (12%) | 809,830 (11%) |
| 5 | 70,366 (6.2%) | 427,239 (7.1%) | 497,605 (6.9%) |
| 6 | 45,042 (4.0%) | 242,827 (4.0%) | 287,869 (4.0%) |
| 7 | 28,209 (2.5%) | 133,834 (2.2%) | 162,043 (2.3%) |
| >= 8 | 46,628 (4.1%) | 171,541 (2.8%) | 218,169 (3.0%) |
| Distance in Miles to Closest Ties |  |  |  |
| Less than 25 miles | 1,034,774 (95%) | 5,462,018 (94%) | 6,496,792 (94%) |
| 25 - 100 miles | 18,922 (1.7%) | 114,161 (2.0%) | 133,083 (1.9%) |
| 100 - 500 miles | 22,662 (2.1%) | 109,821 (1.9%) | 132,483 (1.9%) |
| Over 500 miles | 16,776 (1.5%) | 114,255 (2.0%) | 131,031 (1.9%) |
| Household Annual Income Range (USD\$) | | | |
| 0 - 25,000 | 128,351 (11%) | 118,164 (2.0%) | 246,515 (3.4%) |
| 25,001 - 30,000 | 70,291 (6.2%) | 108,019 (1.8%) | 178,310 (2.5%) |
| 30,001 - 35,000 | 93,639 (8.3%) | 175,951 (2.9%) | 269,590 (3.8%) |
| 35,001 - 40,000 | 112,300 (9.9%) | 271,039 (4.5%) | 383,339 (5.3%) |
| 40,001 - 50,000 | 250,880 (22%) | 992,696 (16%) | 1,243,576 (17%) |
| 50,001 - 60,000 | 187,802 (17%) | 1,263,529 (21%) | 1,451,331 (20%) |
| 60,001 - 80,000 | 220,240 (19%) | 1,760,367 (29%) | 1,980,607 (28%) |

| Table 2 continued from previous page |  |  |  |
| --- | --- | --- | --- |
| Characteristic | Black*,<br>N = 1,253,042<sup>1</sup> | Not Black*,<br>N = 6,828,038<sup>1</sup> | Overall,<br>N = 8,081,080<sup>1</sup> |
| 80,001 - 100,000 | 62,133 (5.5%) | 1,078,729 (18%) | 1,140,862 (16%) |
| 100,001 - 150,000+ | 6,800 (0.6%) | 271,404 (4.5%) | 278,204 (3.9%) |
| Elixhauser Comorbidity Index Score** |  |  |  |
| 0 - <11 | 1,078,365 (86%) | 5,468,567 (80%) | 6,546,932 (81%) |
| 11 - 15 | 80,479 (6.4%) | 578,068 (8.5%) | 658,547 (8.1%) |
| >15 | 94,165 (7.5%) | 781,245 (11%) | 875,410 (11%) |
| Hyperkalemia (Yes) | 4,833 (0.4%) | 20,583 (0.3%) | 25,416 (0.3%) |
| CKD Status |  |  |  |
| CKD Using 2009 | 1,797 (0.1%) | 16,048 (0.2%) | 17,845 (0.2%) |
| CKD Using 2021 | 62,034 (5.0%) | 1,471 (<0.1%) | 63,505 (0.8%) |
| CKD Using Both | 158,640 (13%) | 999,195 (15%) | 1,157,835 (14%) |
| No CKD | 1,030,538 (82%) | 5,811,166 (85%) | 6,841,704 (85%) |
| Did Stage Increase or Decrease with 2021 CKD-EPI Equation? |  |  |  |
| Decreased | 6,461 (4.1%) | 15,394 (1.5%) | 21,855 (1.9%) |
| Stayed the Same | 116,648 (74%) | 981,090 (98%) | 1,097,738 (95%) |
| Increased | 35,531 (22%) | 2,711 (0.3%) | 38,242 (3.3%) |
| Timing of Diagnosis |  |  |  |
| First with 2009 Equation | 9,281 (4.2%) | 30,150 (3.0%) | 39,431 (3.2%) |
| First with 2021 Equation | 53,848 (24%) | 6,901 (0.7%) | 60,749 (4.9%) |
| No Discordance | 159,342 (72%) | 979,663 (96%) | 1,139,005 (92%) |
| *Black refers to Black or African American Individuals |  |  |  |
| **Elixhauser Comorbidity Index computed using van Walraven weights. |  |  |  |
| Categories correlate to low, medium, and high comorbidity. |  |  |  |
| <sup>1</sup>n (%); Median (IQR) |  |  |  |

### Discordance of CKD Staging

In Table 3, we see the discordance of staging identified using the two formulas. 15% of all patients with serum creatinine laboratory values were categorized as having CKD stage 3a, 3b, 4, or 5 using either equation and are included in this sub-analysis. For example, patients that met the criteria for stage 3a, 3b, 4, or 5 using the 2009 CKD-EPI creatinine equation but not the 2021 CKD-EPI creatinine equation are found in the column “No CKD, Stage 1 or 2”.

**Table 3:**
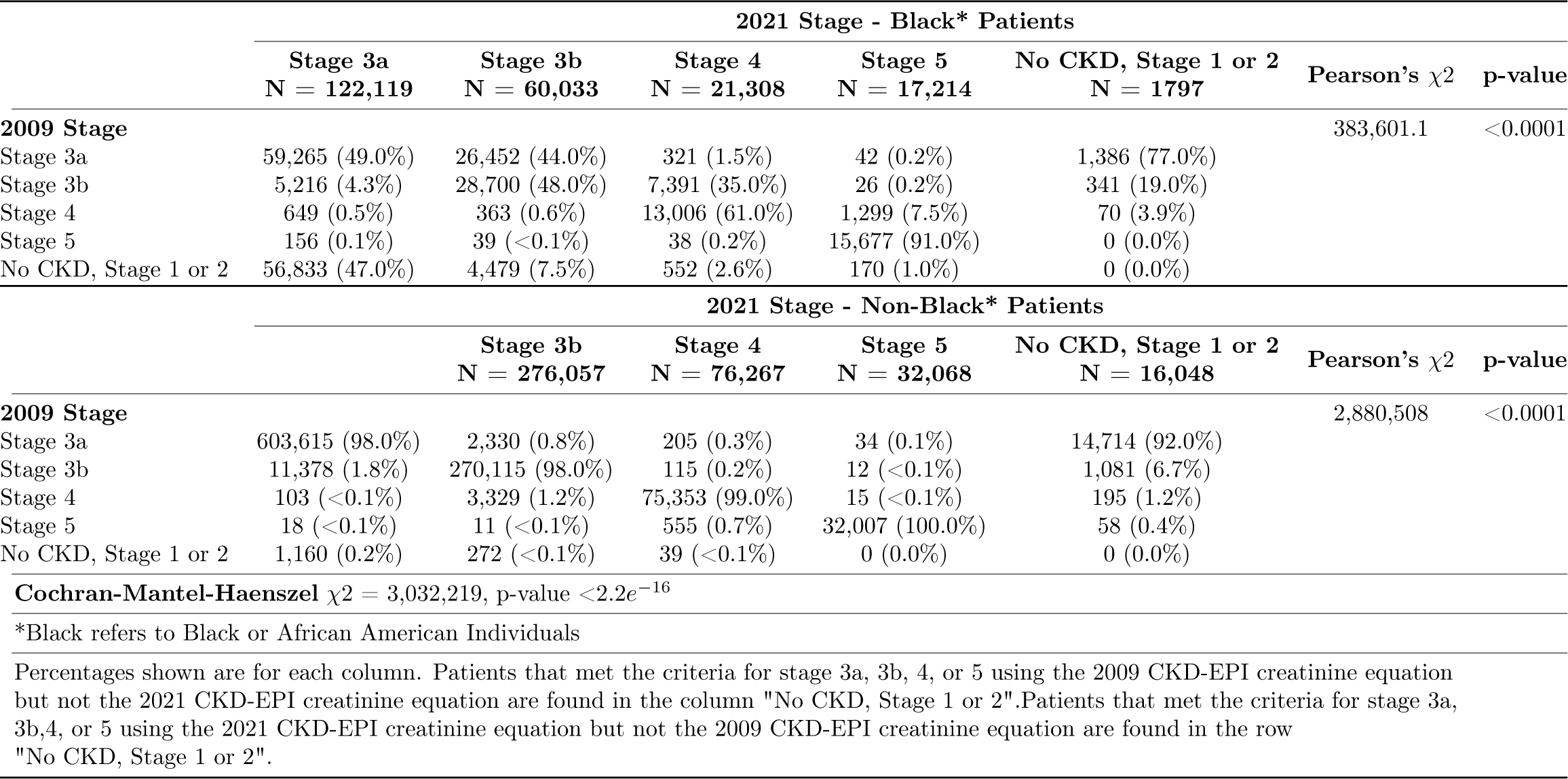
Discordance of Initial Staging for Chronic Kidney Disease by Race Using the 2009 and 2021 CKD-EPI GFR creatinine equations.

Within Black and non-Black patients, the staging category represents patients with concordant diagnoses - those where the 2009 and 2021 formulas yield the same stage. Overall, we found significant discordance in the staging of CKD by equation. Most discordance in the staging of CKD is in the earlier stages of the disease (3a and 3b) than in end stage renal disease (stage 5). Nearly 50% of Black patients that met the criteria for stage 3a using the 2021 equation were found to not meet the criteria for CKD using the 2009 equation. 50% of Black patients that met the criteria for stage 3b using the 2021 equation were categorized as stage 3a using the 2009 equation. Furthermore, approximately 40% of stage 4 disease for Black patients was discordant between the two equations; approximately 36.5% of these patients fell into stage 3a or 3b and 2.6% met the criteria to be stage 5 using the 2009 equation. In contrast, 99% of non-Black patients that were found to be stage 4 using the 2021 equation also were found to be stage 4 using the 2009 equation.

### Multivariate analysis

The final logistic regression model (Table 4 included all covariates as the full model had the lowest AIC. Having a lower number of comorbidities was associated with an increase in staging compared to people with a high comorbidity score, though the difference is small (OR 1.05, CI 1.01, 1.09). Compared to patients that were staged the same (non-discordant) using both equations, having CKD identified earlier in time using the 2009 equation was significantly associated with having a higher stage when using the 2021 equation (OR 5.23, CI 4.97, 5.50). In other words, when a patient was identified as having CKD earlier with the 2021 equation, the patient had a significantly lower chance of being diagnosed at a higher stage of CKD after adjusting for all other factors in the model. Finally, Black patients were 126 times more likely to have an increase in their stage of CKD using the 2021 equation than their counterparts (CI 120, 133).

**Table 4:**
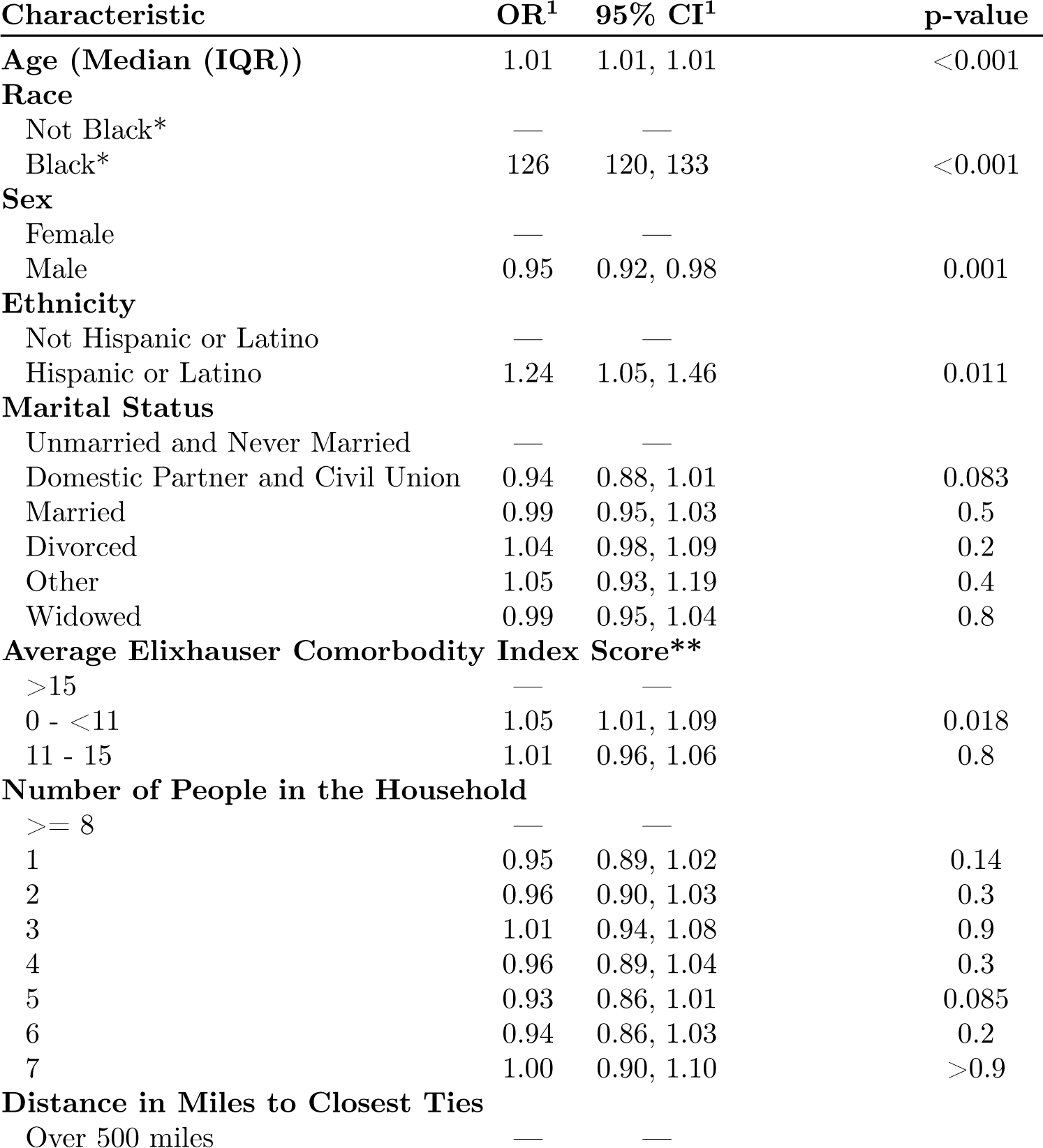
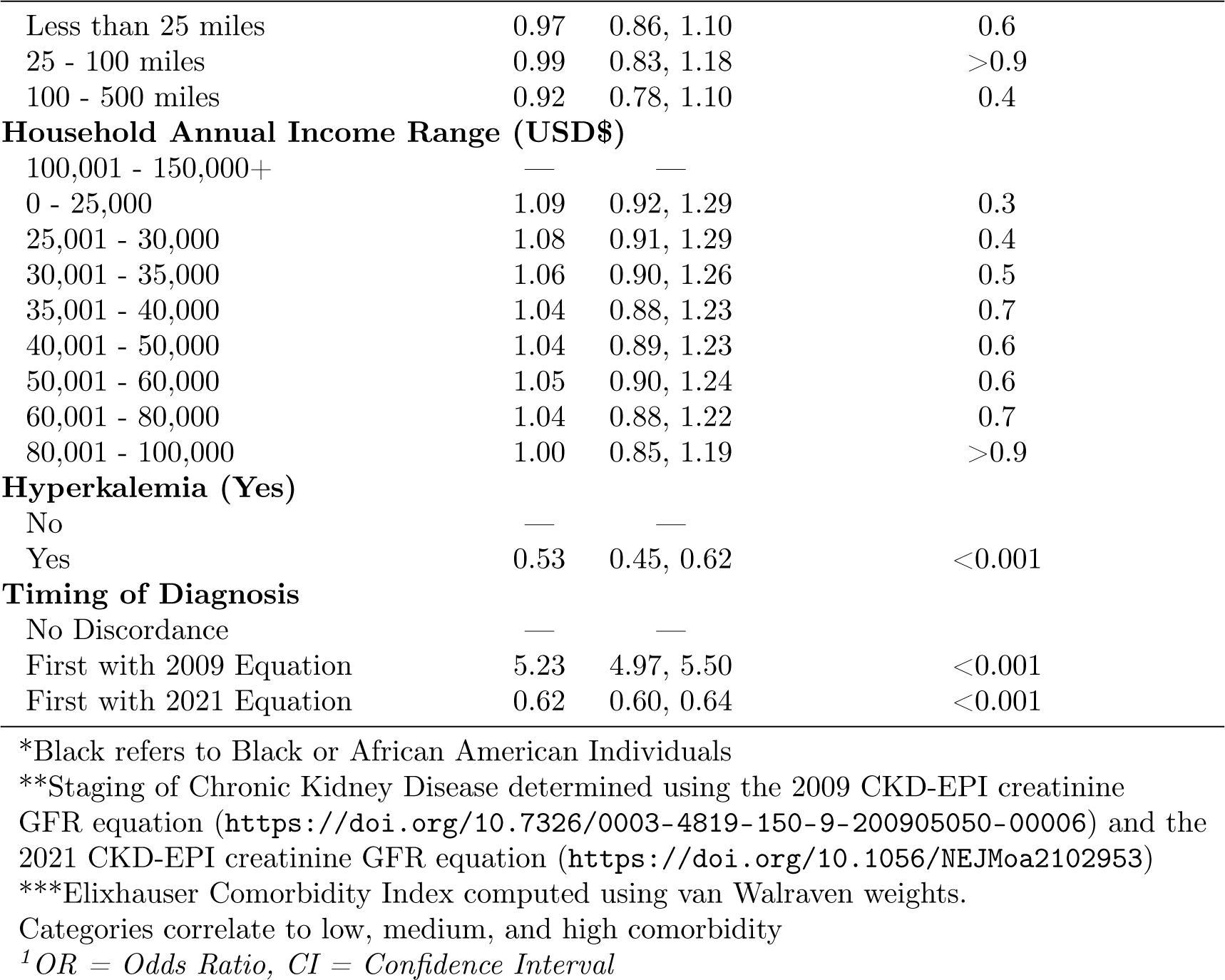
Impact of Demographics, Social Determinants and Comorbidities on the Discordance of Staging of Chronic Kidney Disease**.

| Characteristic | OR<sup>1</sup> | 95% CI<sup>1</sup> | p-value |
| --- | --- | --- | --- |
| **Age (Median (IQR))** | 1.01 | 1.01, 1.01 | <0.001 |
| **Race** |  |  |  |
| Not Black* | — | — |  |
| Black* | 126 | 120, 133 | <0.001 |
| **Sex** |  |  |  |
| Female | — | — |  |
| Male | 0.95 | 0.92, 0.98 | 0.001 |
| **Ethnicity** |  |  |  |
| Not Hispanic or Latino | — | — |  |
| Hispanic or Latino | 1.24 | 1.05, 1.46 | 0.011 |
| **Marital Status** |  |  |  |
| Unmarried and Never Married | — | — |  |
| Domestic Partner and Civil Union | 0.94 | 0.88, 1.01 | 0.083 |
| Married | 0.99 | 0.95, 1.03 | 0.5 |
| Divorced | 1.04 | 0.98, 1.09 | 0.2 |
| Other | 1.05 | 0.93, 1.19 | 0.4 |
| Widowed | 0.99 | 0.95, 1.04 | 0.8 |
| **Average Elixhauser Comorbidity Index Score\*\*** |  |  |  |
| >15 | — | — |  |
| 0 - <11 | 1.05 | 1.01, 1.09 | 0.018 |
| 11 - 15 | 1.01 | 0.96, 1.06 | 0.8 |
| **Number of People in the Household** |  |  |  |
| >= 8 | — | — |  |
| 1 | 0.95 | 0.89, 1.02 | 0.14 |
| 2 | 0.96 | 0.90, 1.03 | 0.3 |
| 3 | 1.01 | 0.94, 1.08 | 0.9 |
| 4 | 0.96 | 0.89, 1.04 | 0.3 |
| 5 | 0.93 | 0.86, 1.01 | 0.085 |
| 6 | 0.94 | 0.86, 1.03 | 0.2 |
| 7 | 1.00 | 0.90, 1.10 | >0.9 |
| **Distance in Miles to Closest Ties** |  |  |  |
| Over 500 miles | — | — |  |
| Less than 25 miles | 0.97 | 0.86, 1.10 | 0.6 |
| 25 - 100 miles | 0.99 | 0.83, 1.18 | >0.9 |
| 100 - 500 miles | 0.92 | 0.78, 1.10 | 0.4 |
| **Household Annual Income Range (USD\$)** | | | |
| 100,001 - 150,000+ | — | — |  |
| 0 - 25,000 | 1.09 | 0.92, 1.29 | 0.3 |
| 25,001 - 30,000 | 1.08 | 0.91, 1.29 | 0.4 |
| 30,001 - 35,000 | 1.06 | 0.90, 1.26 | 0.5 |
| 35,001 - 40,000 | 1.04 | 0.88, 1.23 | 0.7 |
| 40,001 - 50,000 | 1.04 | 0.89, 1.23 | 0.6 |
| 50,001 - 60,000 | 1.05 | 0.90, 1.24 | 0.6 |
| 60,001 - 80,000 | 1.04 | 0.88, 1.22 | 0.7 |
| 80,001 - 100,000 | 1.00 | 0.85, 1.19 | >0.9 |
| **Hyperkalemia (Yes)** |  |  |  |
| No | — | — |  |
| Yes | 0.53 | 0.45, 0.62 | <0.001 |
| **Timing of Diagnosis** |  |  |  |
| No Discordance | — | — |  |
| First with 2009 Equation | 5.23 | 4.97, 5.50 | <0.001 |
| First with 2021 Equation | 0.62 | 0.60, 0.64 | <0.001 |

\*Black refers to Black or African American Individuals
\*\*Staging of Chronic Kidney Disease determined using the 2009 CKD-EPI creatinine GFR equation (https://doi.org/10.7326/0003-4819-150-9-200905050-00006) and the 2021 CKD-EPI creatinine GFR equation (https://doi.org/10.1056/NEJMoa2102953)
\*\*\*Elixhauser Comorbidity Index computed using van Walraven weights.
Categories correlate to low, medium, and high comorbidity
<sup>1</sup>OR = Odds Ratio, CI = Confidence Interval

## Discussion

Well established medical algorithms with race components are being re-examined in the wake of the broader medical community acceptance that race is a social not biological construct.^46, 11^ The inclusion of race in many of these algorithms goes back decades if not centuries as vestiges of race science.^47^ The longstanding method of diagnosing CKD is no exception. There are multiple equations to estimate the GFR, but the Kidney Disease: Improving Global Outcomes (KDIGO) organization recommends calculating it using serum creatinine. The 2021 update from the CKD-EPI task force removed the race adjustment^9^ and works have been published both lauding the change and arguing that race should not be excluded from the determination. Work in countries that consider their populations to have a more homogenous racial makeup found significant negative changes in the prevalence of CKD when using the 2021 CKD-EPI equation and conclude that the equation is biased.^48, 49, 50^ These works in particular do not support that race should be included when estimating GFR (one in particular did not include the race coefficient when using the 2009 equation^48^) but do support that the equation potentially has other errors that do not appropriately estimate kidney function. Additionally, works to validate the change have been implemented and used as evidence to support the positivity of using the 2021 CKD-EPI equation.^51, 9, 52, 10, 53^

The present study aimed to understand whether using the 2021 CKD-EPI equation led to decreased disparity between Black patients and their counterparts with regards to how quickly they should have been diagnosed with CKD. We also aimed to find whether there was discordance in the staging at diagnosis for Black patients compared to their counterparts. Given the attention this topic has sustained, our goal was to add evidence from a large diverse national cohort using EHR data while including some important SDOH factors such as closeness to social network and overall household income. In our study of over 8 million patients, we found that the 2021 CKD-EPI equation did result in earlier identification of CKD (GFR < 60) for 24% of Black patients but only 0.7% of non-Black patients. Initial staging of the disease is where more of the discrepancies were found, especially among Black patients. The disconnect in the timing of staging for Black patients means a number of these patients may not have received the appropriate treatment and care they needed including dialysis or transplants due to being diagnosed with a later stage at a later time than what was found with the 2021 CKD-EPI equation. This discordance in staging may have contributed to earlier morbidity or mortality for this population.^54, 55, 16^

The 2021 CKD-EPI GFR calculation still includes adjustments for sex which may be a major contributor to discrepancies in both diagnosis and staging of the disease. As recognized from many epidemiological studies, women are far more likely to obtain medical care than men and obtain care more often.^13^ Finding out the amount of influence being female would change the estimation of GFR is beyond the scope of this study.

We included several SDOH factors in this study that were previously identified as being related to CKD.^51, 56^ These previous works highlighted groups that should be targeted for improved care such as people who are poor (near or below the federal poverty level) or who live in poor areas. In this study, we did not find that any of our included SDOH factors were significantly associated with having an increased stage of CKD when identified with the 2021 CKD-EPI equation. This indicates that more effort is needed to identify more specific and available SDOH factors in EHR data for future work such as the inclusion of the social vulnerability index or poverty level data.

Prevention of CKD and early intervention can reduce the need for or improve the outcomes of downstream care needs such as dialysis or transplantation. We included distance to close relatives in terms of miles and how many people were in a household, but these were not statistically significant factors in the present study. Due to other research, we do know that factors such as closeness to a social network directly impact both the diagnosis of conditions [friends or partners influence the seeking of medical care] but also the treatment.^57, 56^ A study conducted on transplant donors found that more than 25% of Black donors who were not diagnosed with CKD at the time of their donation with the formulas used then would today be reclassified as having CKD.^58^ The effect of this statistic on the disparity of poor outcomes after transplantation for Black patients is unknown but could be substantial. If disease was identified earlier as we identified in the present study, how many known patients could avoid the need for transplantation? If disease was identified earlier as in the present study, how many potential kidney donors would not reduce their own kidney capacity through donation or would be able to access early care as needed? These questions must be addressed.

### Strengths and Limitations

While using a large cohort (8 million patients) is a strength, ours is subject to a variety of known limitations to EHR data. We are only able to identify events that are captured by the constituent health care systems that are a part of the Truveta member system. Events and comorbidity status may be unknown when captured outside of member healthcare systems. These are common and well understood limitations associated with using EHR data.^59^ We were, however, able to view every laboratory result in the data and were not subject to restrictions in only being able to access billing or diagnosis codes. Because of this we were able to directly calculate the exact difference in time of diagnosis.

We used the Elixhauser comorbidity index, a well known tool for including the impact of comorbidity on hospitalization related outcomes, but this method has not been adjusted or validated for this particular population. We also selected to use van Walraven weights with the Elixhauser comorbidity index. This could mean that we over or under estimated the impact of particular comorbid conditions, did not include other comorbid conditions, or that the weights used were not most applicable to this circumstance. We may not have included or excluded all comorbidities that can impact serum creatinine or kidney function.

Despite these limitations, our results still underscore the need to continue to examine the positive impacts of removing race from the estimation of GFR in the US.

## Data Availability

All data used in this study is available to subscribers at studio.truveta.com.

## Acknowledgements

The authors thank the Truveta clinical informatics team for their assistance and input during the design and development of this study.

## Institutional Review Board Approval

Normalized electronic health record data are de-identified by expert determination under the HIPAA Privacy Rule before being made available to researchers. In accordance with 46.101 Protection of Human Subjects, our study did not require Institutional Review Board approval because it used only deidentified medical records. All data used in this study are publicly available to all Truveta subscribers and may be accessed at (studio.truveta.com).

I confirm that all necessary patient/participant consent has been obtained and the appropriate institutional forms have been archived, and that any patient/participant/sample identifiers included were not known to anyone (e.g., hospital staff, patients or participants themselves) outside the research group so cannot be used to identify individuals.

## References

1. Nathan R. Hill, Samuel T. Fatoba, Jason L. Oke, Jennifer A. Hirst, Christopher A. O’Callaghan, Daniel S. Lasserson, and F. D. Richard Hobbs. Global prevalence of chronic kidney disease – a systematic review and meta-analysis. PLOS ONE, 11(7):1–18, 07 2016.

2. Marcello Tonelli, Natasha Wiebe, Bruce Guthrie, Matthew T. James, Hude Quan, Martin Fortin, Scott W. Klarenbach, Peter Sargious, Sharon Straus, Richard Lewanczuk, Paul E. Ronksley, Braden J. Manns, and Brenda R. Hemmelgarn. Comorbidity as a driver of adverse outcomes in people with chronic kidney disease. Kidney International, 2015.

3. Keith Norris and Allen R. Nissenson. Race, gender, and socioeconomic disparities in ckd in the united states. Journal of the American Society of Nephrology, 2008.

4. Centers for Disease Control and Prevention. Chronic kidney disease in the united states, 2023. https://www.cdc.gov/kidneydisease/publications-resources/CKD-national-facts.htm l, 2023.

5. Claudia S. Walker and Crystal A. Gadegbeku. Addressing kidney health disparities with new national policy: the time is now. Cardiovascular Diagnosis and Therapy, 2023.

6 National Kidney Foundation. Health disparities | national kidney foundation. https://www.kidney.org/advocacy/legislative-priorities/health-disparities.

7. Susan E. Quaggin and Paul M. Palevsky. Removing race from kidney disease diagnosis. Journal of the American Society of Nephrology, 2021.

8. Andrew S. Levey, Silvia M. Titan, Neil R. Powe, Josef Coresh, and Lesley A. Inker. Kidney disease, race, and gfr estimation. Clinical Journal of the American Society of Nephrology, 2020.

9. Cynthia Delgado, Mukta Baweja, Deidra C Crews, Nwamaka D Eneanya, Crystal A Gadegbeku, Lesley A Inker, Mallika L Mendu, W Greg Miller, Marva M Moxey-Mims, Glenda V Roberts, Wendy L St. Peter, Curtis Warfield, and Neil R Powe. A unifying approach for gfr estimation: Recommendations of the nkf-asn task force on reassessing the inclusion of race in diagnosing kidney disease. Journal of the American Society of Nephrology, 32:2994, 2021.

10. Prabhdeep Uppal, Benjamin L Golden, Ashley Panicker, Omar A Khan, and Matthew J Burday. The case against race-based gfr. Delaware journal of public health, 8:86–89, 8 2022.

11. Bridget M. Kuehn. Medical students lead effort to remove race from kidney function estimates. Kidney News, 12(7):1 – 3, 2020.

12. Neil R Powe. Black kidney function matters: Use or misuse of race? JAMA, 324:737–738, 2020.

13. Lesley A. Inker, Nwamaka D. Eneanya, Josef Coresh, Hocine Tighiouart, Dan Wang, Yingying Sang, Deidra C. Crews, Alessandro Doria, Michelle M. Estrella, Marc Froissart, Morgan E. Grams, Tom Greene, Anders Grubb, Vilmundur Gudnason, Orlando M. Gutiérrez, Roberto Kalil, Amy B. Karger, Michael Mauer, Gerjan Navis, Robert G. Nelson, Emilio D. Poggio, Roger Rodby, Peter Rossing, Andrew D. Rule, Elizabeth Selvin, Jesse C. Seegmiller, Michael G. Shlipak, Vicente E. Torres, Wei Yang, Shoshana H. Ballew, Sara J. Couture, Neil R. Powe, and Andrew S. Levey. New creatinine-and cystatin c–based equations to estimate gfr without race. New England Journal of Medicine, 385(19):1737–1749, 2021. PMID: 34554658.

14. Pierre Delanaye, Ingrid Masson, Nicolas Maillard, Hans Pottel, and Christophe Mariat. The new 2021 ckd-epi equation without race in a european cohort of renal transplanted patients. Transplantation, 2022.

15. Orlando M. Gutiérrez, Yingying Sang, Morgan E. Grams, Shoshana H. Ballew, Aditya Surapaneni, Kunihiro Matsushita, Alan S. Go, Michael G. Shlipak, Lesley A. Inker, Nwamaka D. Eneanya, Deidra C. Crews, Neil R. Powe, Andrew S. Levey, Josef Coresh, and Chronic Kidney Disease Prognosis Consortium. Association of Estimated GFR Calculated Using Race-Free Equations With Kidney Failure and Mortality by Black vs Non-Black Race. JAMA, 327(23):2306–2316, 06 2022.

16. Ebele M. Umeukeje, Taneya Y. Koonce, Sheila V. Kusnoor, Ifeoma I. Ulasi, Sophia Kostelanetz, Annette M. Williams, Mallory N. Blasingame, Marcia I. Epelbaum, Dario A. Giuse, Annie N. Apple, Karampreet Kaur, Tavia González Peña, Danika Barry, Leo G. Eisenstein, Cameron T. Nutt, and Nunzia B. Giuse. Systematic review of international studies evaluating mdrd and ckdepi estimated glomerular filtration rate (egfr) equations in black adults. PLOS ONE, 17(10):1–17, 10 2022.

17. James A. Diao, Gloria J. Wu, Jason K. Wang, Isaac S. Kohane, Herman A. Taylor, Hocine Tighiouart, Andrew S. Levey, Lesley A. Inker, Neil R. Powe, and Arjun K. Manrai. National projections for clinical implications of race-free creatinine-based gfr estimating equations. Journal of the American Society of Nephrology, 34:309–321, 2 2023.

18. Andrew S. Levey, Lesley A. Stevens, Christopher H. Schmid, Yaping (Lucy) Zhang, Alejandro F. Castro, Harold I. Feldman, John W. Kusek, Paul Eggers, Frederick Van Lente, Tom Greene, and Josef Coresh and. A new equation to estimate glomerular filtration rate. Annals of Internal Medicine, 2009.

19. A Elixhauser, C Steiner, D R Harris, and R M Coffey. Comorbidity measures for use with administrative data. Medical Care, 36:8–27, 1998.

20. Carl Van Walraven, Peter C. Austin, Alison Jennings, Hude Quan, and Alan J. Forster. A modification of the elixhauser comorbidity measures into a point system for hospital death using administrative data. Medical care, 47:626–633, 6 2009.

21. Juliëtte F. Velu and Jan Baan Jr. Elixhauser comorbidity score is the best risk score in predicting survival after mitraclip implantation. Structural Heart, 2(1):53–57, 2018.

22 National Kidney Foundation. How to classify ckd | national kidney foundation. https://www.kidney.org/professionals/explore-your-knowledge/how-to-classify-ckd.

23 Python Software Foundation. python, June 2022.

24 Guido Van Rossum and Fred L. Drake. Python 3 Reference Manual. CreateSpace, Scotts Valley, CA, 2009.

25 Wes McKinney. Data Structures for Statistical Computing in Python. In Stéfan van der Walt and Jarrod Millman, editors, Proceedings of the 9th Python in Science Conference, pages 56–61, 2010.

26 The pandas development team. pandas-dev/pandas: Pandas. 10.5281/zeno do.7549438, jan 2023.

27. Charles R. Harris, K. Jarrod Millman, Stéfan J. van der Walt, Ralf Gommers, Pauli Virtanen, David Cournapeau, Eric Wieser, Julian Taylor, Sebastian Berg, Nathaniel J. Smith, Robert Kern, Matti Picus, Stephan Hoyer, Marten H. van Kerkwijk, Matthew Brett, Allan Haldane, Jaime Fernández del Río, Mark Wiebe, Pearu Peterson, Pierre Gérard-Marchant, Kevin Sheppard, Tyler Reddy, Warren Weckesser, Hameer Abbasi, Christoph Gohlke, and Travis E. Oliphant. Array programming with NumPy. Nature, 585(7825):357–362, September 2020.

28. Kirill Simonov. PyYAML, October 2021.

29. J. D. Hunter. Matplotlib: A 2d graphics environment. Computing in Science & Engineering, 9(3):90–95, 2007.

30 R Core Team. R: A Language and Environment for Statistical Computing. Vienna, Austria, 2022. https://www.R-project.org/.

31. Hadley Wickham, Romain François, Lionel Henry, and Kirill Müller. dplyr: A Grammar of Data Manipulation, 2022. https://CRAN.R-project.org/package=dplyr.

32. Hadley Wickham, Romain François, Lionel Henry, Kirill Müller, and Davis Vaughan. dplyr: A Grammar of Data Manipulation, 2023. R package version 1.1.3.

33 Hadley Wickham. ggplot2: Elegant Graphics for Data Analysis. Springer-Verlag New York, 2016.

34. Hadley Wickham, Winston Chang, Lionel Henry, Thomas Lin Pedersen, Kohske Takahashi, Claus Wilke, Kara Woo, Hiroaki Yutani, and Dewey Dunnington. ggplot2: Create Elegant Data Visualisations Using the Grammar of Graphics, 2022. R package version 3.4.0.

35 Daniel D. Sjoberg, Joseph Larmarange, Michael Curry, Jessica Lavery, Karissa Whiting, Emily C. Zabor, Xing Bai, Esther Drill, Jessica Flynn, Margie Hannum, Lobaugh, Amy Tin, and Gustavo Zapata Wainberg. gtsummary: Presentation-Ready Data Summary and Analytic Result Tables, January 2023.

36. David Robinson, Alex Hayes, Simon Couch [aut, cre, Posit Software, PBC, Indrajeet Patil, Derek Chiu, Gomez, Dieter Menne, Benjamin Nutter, Johnston, Francois Briatte, Jeffrey Arnold, Gabry, Gavin Simpson, Jens Preussner, Hesselberth, Matthew Lincoln, Gasparini, Frederick Novometsky, Wilson Freitas, Michelle Evans, Jason Cory Brunson, Simon Jackson, Whalley, Yves Rosseel, Michael Kuehn, Cimentada, Karl Dunkle Werner, Ethan Christensen, Steven Pav, Paul PJ, Ben Schneider, Patrick Kennedy, Medina, Jason Muhlenkamp, Matt Lehman, Denney, Andrew Bates, Vincent Arel-Bundock, Hayashi, Annie Wang, Wei Yang Tham, Wang, Jasper Cooper, E. Auden Krauska, Wang, Charles Gray, Jared Wilber, Gegzna, Frederik Aust, Angus Moore, Williams, Bruna Wundervald, Joyce Cahoon, McDermott, Shiro Kuriwaki, Lukas Wallrich, Martherus, Joseph Larmarange, Max Kuhn, Michal Bojanowski, Hakon Malmedal, Clara Wang, Sergio Oller, Sonnet, Ben Schneider, Bernie Gray, Averick, Andreas Bender, Sven Templer, Paul-Christian Buerkner, Matthew Kay, Erwan Le Pennec, Junkka, Benjamin Soltoff, Zoe Wilkinson Saldana, Tyler Littlefield, Charles T. Gray, Shabbh E. Banks, Robinson, Riinu Ots, Nicholas Williams, Jakobsen, Lisa Lendway, Karl Hailperin, Rodriguez, Chris Jarvis, Greg Macfarlane, Mannakee, Shreyas Singh, Laurens Geffert, Ooi, Eduard Szocs, David Hugh-Jones, Matthieu Stigler, Hugo Tavares, R. Willem Vervoort, Wiernik, Jasme Lee, Taren Sanders, Prosdocimi, and Alex Reinhart. broom: Convert Statistical Objects into Tidy Tibbles, March 2023.

37 W. N. Venables and B. D. Ripley. Modern Applied Statistics with S. Springer, New York, fourth edition, 2002.

38. David B. Dahl, David Scott, Charles Roosen, and Magnusson. xtable: Export Tables to LaTeX or HTML. 2019.

39. Hadley Wickham. Reshaping data with the reshape package. Journal of Statistical Software, 21(12):1–20, 2007.

40 Hadley Wickham. reshape2: Flexibly Reshape Data: A Reboot of the Reshape Package, 2020. R package version 1.4.4.

41 Hadley Wickham, Jim Hester, and Jennifer Bryan. readr: Read Rectangular Text Data, 2023. R package version 2.1.4.

42. Neal Richardson, Ian Cook, Nic Crane, Dewey Dunnington, Romain François, Jonathan Keane, Dragos, Moldovan-Grünfeld, Jeroen Ooms, and Apache Arrow. arrow: Integration to Apache ‘Arrow’, 2023. R package version 10.0.1.

43 Hadley Wickham. stringr: Simple, Consistent Wrappers for Common String Operations, 2022. R package version 1.5.0.

44. Javier Luraschi, Kevin Kuo, Kevin Ushey, JJ Allaire, Hossein Falaki, Lu Wang, Andy Zhang, Yitao Li, Edgar Ruiz, and The Apache Software Foundation. sparklyr: R Interface to Apache Spark, 2023. R package version 1.8.2.

45 The Apache Software Foundation. SparkR: R Front End for Apache Spark, 2023. R package version 3.3.1.

46. Andrea Deyrup and Joseph L. Graves. Racial biology and medical misconceptions. New England Journal of Medicine, 386(6):501–503, 2022. PMID: 35119803.

47. Linda M. Chatters, Robert Joseph Taylor, and Amy J. Schulz. The return of race science and why it matters for family science. Journal of Family Theory amp; Review, 2022.

48. Michal J. Lewandowski, Simon Krenn, Amelie Kurnikowski, Philipp Bretschneider, Martina Sattler, Elisabeth Schwaiger, Marlies Antlanger, Philipp Gauckler, Markus Pirklbauer, Maria Brunner, Sabine Horn, Emanuel Zitt, Bernhard Kirsch, Martin Windpessl, Manfred Wallner, Ida Aringer, Martin Wiesholzer, Manfred Hecking, and Sebastian Hödlmoser. Chronic kidney disease is more prevalent among women but more men than women are under nephrological care. Wiener klinische Wochenschrift, 2022.

49. Tae-Dong Jeong, Jinyoung Hong, Woochang Lee, Sail Chun, and Won-Ki Min. Accuracy of the new creatinine-based equations for estimating glomerular filtration rate in koreans. Annals of Laboratory Medicine, 2022.

50. Edouard L Fu, Josef Coresh, Morgan E Grams, Catherine M Clase, Carl-Gustaf Elinder, Julie Paik, Chava L Ramspek, Lesley A Inker, Andrew S Levey, Friedo W Dekker, and Juan J Carrero. Removing race from the ckd-epi equation and its impact on prognosis in a predominantly white european population. Nephrology Dialysis Transplantation, 2022.

51. Jenna M. Norton, Marva M. Moxey-Mims, Paul W. Eggers, Andrew S. Narva, Robert A. Star, Paul L. Kimmel, and Griffin P. Rodgers. Social determinants of racial disparities in ckd. Journal of the American Society of Nephrology, 2016.

52. Cynthia Delgado, Mukta Baweja, Nilka Ríos Burrows, Deidra C Crews, Nwamaka D Eneanya, Crystal A Gadegbeku, Lesley A Inker, Mallika L Mendu, W Greg Miller, Marva M Moxey-Mims, Glenda V Roberts, Wendy L St. Peter, Curtis Warfield, and Neil R Powe. Reassessing the inclusion of race in diagnosing kidney diseases: An interim report from the nkf-asn task force. Journal of the American Society of Nephrology, 32:1305, 2021.

53. James A Diao, Gloria J Wu, Herman A Taylor, John K Tucker, Neil R Powe, Isaac S Kohane, and Arjun K Manrai. Clinical implications of removing race from estimates of kidney function. JAMA, 325:184–186, 2021.

54. Andrew S. Levey, Lesley A. Inker, and Josef Coresh. Gfr estimation: From physiology to public health. American Journal of Kidney Diseases, 2014.

55. Song Lu, Kimberly Robyak, and Yusheng Zhu. The ckd-epi 2021 equation and other creatinine-based race-independent egfr equations in chronic kidney disease diagnosis and staging. The Journal of Applied Laboratory Medicine, 2023.

56. Susanne B. Nicholas, Kamyar Kalantar-Zadeh, and Keith C. Norris. Socioeconomic disparities in chronic kidney disease. Advances in Chronic Kidney Disease, 2015.

57. Deidra C. Crews and Tessa K. Novick. Social determinants of ckd hotspots. Seminars in Nephrology, 2019.

58. Babak J Orandi, Vineeta Kumar, Rhiannon D Reed, Paul A MacLennan, Brittany A Shelton, Chandler McLeod, and Jayme E Locke. Reclassification of ckd in living kidney donors with the refitted race-free egfr formula. The American Journal of Surgery, 10 2022. doi: 10.1016/j.amjsurg.2022.09.024.

59. Suchitra Kataria and Vinod Ravindran. Electronic health records: A critical appraisal of strengths and limitations. Journal of the Royal College of Physicians of Edinburgh, 50(3):262–268, sep 2020.

